# Genome-wide DNA methylation-analysis delineates blastic plasmacytoid dendritic cell neoplasm from related entities and identifies distinct molecular features

**DOI:** 10.1101/2023.07.28.23293273

**Authors:** Axel Künstner, Julian Schwarting, Hanno M. Witte, Pengwei Xing, Veronica Bernard, Stephanie Stölting, Philipp Lohneis, Florian Janke, Maede Salehi, Xingqi Chen, Kathrin Kusch, Holger Sültmann, Emil Chteinberg, Anja Fischer, Reiner Siebert, Nikolas von Bubnoff, Hartmut Merz, Hauke Busch, Alfred C. Feller, Niklas Gebauer

**Author notes:** These authors contributed equally to this manuscript (Axel Künstner, Julian Schwarting, Hanno M. Witte). Shared senior authorship (Hauke Busch, Alfred C. Feller, Niklas Gebauer). **Corresponding Author:** PD Dr. med. Niklas Gebauer Department of Hematology and Oncology UKSH Campus Luebeck Ratzeburger Allee 160 23538 Luebeck.

## Abstract

Blastic plasmacytoid dendritic cell neoplasm (BPDCN) constitutes a rare and aggressive malignancy originating from plasmacytoid/common dendritic cells (pDCs/cDCs) with a primarily cutaneous tropism followed by dissemination to the bone marrow and other organs. We conducted a genome-wide analysis of the tumor methylome in an extended cohort of 45 BPDCN patients supplemented by WES (n=54) and RNA-seq (n=54) as well as ATAC-seq on selected cases (n=4). We determine the BPDCN DNA methylation profile and thereby identify a reliable means to discriminate BPDCN from AML, CMML and T-ALL. DNA methylation profiling characterizes disruption of oncogenic pathways whilst unraveling the proliferative history as well as the prognostically relevant composition of the tumor microenvironment. Beyond the two recently established BPDCN subtypes (C1/C2), we identified a transcriptional reliance on JAK/STAT and NFκB-signaling in atypical C2 versus C1-BPDCN cases through RNA-sequencing. Our integrative characterization of BPDCN offers novel molecular insights and potential diagnostic applications.

## Introduction

Blastic plasmacytoid dendritic cell neoplasm (BPDCN) is an aggressive and extremely rare blood cancer, accounting for approx. 0.5% of acute hematological malignancies. In its recent editions, the WHO classification of myeloid neoplasms recognizes BPDCN as a distinct entity descending from non-activated, CD56^+^ plasmacytoid dendritic cells (pDC) ^1–3^. However, a broader cellular origin encompassing transcriptional signatures of both AXL1^+^ SIGLEC6^+^ DCs and earlier, common dendritic cells, termed AS-DCs and cDCs, respectively, has been proposed, before, suggesting a diverse cellular ontogeny ^4–6^. Clinically, skin lesions commonly precede bone marrow infiltration and secondary propagation into lymph nodes and extranodal organs. A striking 4:1 male predominance, attributed to sex-biased *ZRSR2* mutations and enrichment in elderly patients with a median age of around 70 years at diagnosis has been observed ^7, 8^. While the typical BPDCN immunophenotype (CD4^+^, CD56^+^, CD123^+^) is relatively specific and reliably enables correct diagnosis, discrimination from AML, especially cases with pDC features can be challenging ^9, 10^. Investigation of pDC– associated antigens (e.g., TCL1 or CD303) can further facilitate differential diagnosis, while the expression of B-(CD79b), T-(CD2, CD7) and precursor antigens (Tdt) poses variable pitfalls ^11^. Treatment with conventional chemotherapy alone results in insufficient and short-lived remissions (median overall survival (OS) of 12 to 14 months), which necessitates either allogeneic or autologous stem cell transplantation (SCT) in therapeutic approaches of curative intent ^7, 12, 13^. Only through the introduction of tagraxofusp, a CD123-directed cytotoxin, which recently demonstrated high clinical efficacy, curative treatment has become possible in elderly patients, as well, while the outcome in relapsed/refractory cases remains dismal ^14^.

Genome-wide DNA methylation profiling has evolved from a descriptive, ontological analysis into a diagnostic assay of prognostic relevance across a variety of solid cancers, including CNS tumors and sarcomas ^15, 16^. While genomic and transcriptional profiling has revolutionized the taxonomy of myeloid cancers, embedded in the current WHO classification, the study of DNA methylation may add another layer of insight into BPDCN biology that is so far insufficiently captured by DNA- and RNA-sequencing ^1^. Methylome analysis may further assist in differential diagnostics between BPDCN and other malignancies sharing close molecular ties including chronic myelomonocytic leukemia (CMML), acute myeloid leukemia (AML) and myelodysplastic syndromes (MDS), which may occur syn- and metachronously in up to 20% of cases ^17^.

DNA methylation profiling can further assess the tumor immune microenvironment (TME), by deconvoluting cell types from within tissue-derived bulk DNA and discriminating immunologically hot from cold tumors ^18^. The robust correlation of its output with an immunohistochemical dissection of the TME has been shown in non-small cell lung cancer and others ^18, 19^.

In this study, we have extended our previously published BPDCN cohort^6^, assessed by paired whole-exome (WES) and RNA-sequencing (RNA-seq) as well as genome-wide copy number analysis and conducted array-based genome-wide DNA methylation profiling and ATAC-sequencing on selected cases, allowing for a more profound and novel understanding of BPDCN pathobiology and reliable discrimination from AML as the predominant diagnostic challenge in clinical practice. Moreover, we identify two immunological subtypes, characterized by features in the TME and recurrent genomic alterations.

## Results

### Clinical characteristics of the study group and expanded deconvolution cohort

We collected 54 quality controlled (diagnostic BPDCN samples with sufficient formalin-fixed paraffin-embedded (FFPE) tissues (age range 15 – 91 years; mean/median 69/72 years). We observed an expectedly strong male predominance (40/54; 74%) and uniform cutaneous involvement alongside a pronounced proclivity for extranodal involvement. Of patients in first complete (CR) or partial remission (PR), 24% went on to autologous/allogeneic SCT. Clinical outcomes, when available, reflected previous dismal observations in BPDCN with a median progression-free and overall survival of eight and twelve months, respectively. Best supportive care was provided in four cases where any form of anti-neoplastic treatment was refused.

BPDCN-specific markers were expressed in all cases, whereas precursor antigens (CD34, TdT) were present in 5/47 (11%) and 34/47 (72%) respectively. Baseline clinicopathological characteristics of BPDCN cases included in the current study are briefly summarized in **Table 1**. In accordance with our previous study, RNA-seq data were deconvoluted according to single-cell DC and monocyte datasets and previous observations were recapitulated/extended to new samples including differentially mutated genes (**Supplementary Figures 1 and 2**).

**Table 1.** Baseline clinicopathological characteristics of the study group.

| Characteristics | BPDCN-C1<br>(n = 29) | BPDCN-C2<br>(n = 25) |
| --- | --- | --- |
| Age (yrs.; median (range)) | 70 (15 - 91) | 74 (42 – 90) |
| **Sex** |  |  |
| Female | 6 (21%) | 8 (32%) |
| Male | 23 (79%) | 17 (68%) |
| **Manifestation** |  |  |
| Skin | 20 (69%) | 14 (56%) |
| Bone marrow | 8 (28%) | 8 (32%) |
| 0 EN-sites | 3 (10%) | 3 (12%) |
| 1-2 EN sites | 26 (90%) | 19 (76%) |
| > 2 EN sites | - | 3 (12%) |
| **Stage (Ann Arbor)** |  |  |
| I/II | 2/16 (13%) | 3/16 (19%) |
| III/IV | 14/16 (87%) | 13/16 (81%) |
| **ECOG PS** |  |  |
| 0-1 | 8/10 (80%) | 3/11 (27%) |
| ≥2 | 2/10 (20%) | 8/11 (73%) |
| **B-symptoms** |  |  |
| No | 6/14 (43%) | 4/15 (27%) |
| Yes | 8/14 (57%) | 11/15 (73%) |
| **Immunohistochemistry** |  |  |
| BPDCN-specific | 29/29 (100%) | 25/25 (100%) |
| CD56<sup>+</sup> | 29/29 (100%) | 25/25 (100%) |
| CD123<sup>+</sup> | 29/29 (100%) | 25/25 (100%) |
| TCL1 | 27/28 (96%) | 17/23 (74%) |
| Immature lineage marker | 21/29 (72%) | 19/24 (79%) |
| CD34 | 1/29 (3%) | 4/24 (17%) |
| TdT<sup>+</sup> | 21/29 (72%) | 16/23 (70%) |
| T-lineage markers | 28/29 (96%) | 24/25 (96%) |
| CD2<sup>+</sup> | 5/22 (23%) | 7/16 (44%) |
| CD3<sup>+</sup> | 4/26 (15%) | 5/23 (22%) |
| CD4<sup>+</sup> | 28/29 (96%) | 23/25 (92%) |
| B-lineage marker CD79A<sup>+</sup> | 23/28 (82%) | 20/23 (87%) |
| Myeloid-lineage markers | 24/26 (92%) | 21/23 (91%) |
| CD33<sup>+</sup> | 24/26 (92%) | 20/23 (87%) |
| CD117<sup>+</sup> | 2/26 (8%) | 9/23 (39%) |
| MPO<sup>+</sup> | 1*/28 (4%) | 4*/23 (17%) |
| Ki-67 (median, range) | 60% (30 – 90%) | 50% (25 – 90%) |
| Abbreviations: BPDCN, blastic plasmacytoid dendritic cell neoplasm; ECOG, Eastern Cooperative Oncology Group; EN, extranodal; MPO, myeloperoxidase; yrs, years. *cases with concurrent other myeloid neoplasia (CMML or AML). |  |  |

Additionally, mutational landscape and MutSigCV analysis were extended to new cases as previously described (**Supplementary Tables 1 and 2**) ^6^. Tumor mutational burden (TMB) was confirmed to be significantly higher (p = 0.0055) among C1-BPDCN and an updated set of differentially mutated genes was identified (**Supplementary Figures 3a and 3b**).

### Epigenetic profiling reveals significant deregulation of key regulatory pathways through loss of DNA methylation compared to dendritic cells

In order to assess epigenetic processes contributing to the malignant transformation from DCs to BPDCN and to allocate the entity within the spectrum of blood cells, we performed a principal component analysis (PCA) of BPDCN and various cell types of the peripheral blood (**Figure 1**) ^20^. Expectedly, we observed a clear segregation of blood cell types and BPDCN, but beyond this, a marked difference between BPDCN and previously profiled DC subsets became apparent (**Figure 1a**) ^21^. This was further reflected in significantly higher global mean DNA methylation levels in both early and more mature DCs, signifying the dramatic extent of DNA methylation loss during malignant transformation of DCs towards BPDCN (**Figure 1b**). An enrichment analysis of differentially methylated regions (CpG FDRs < 0.01, absolute difference above 0.3, enrichment analysis against HALLMARK and REACTOME gene sets) revealed significantly reduced DNA methylation in candidate genes involved in processes like extracellular matrix organization, collagen modulation and the neuronal system as recently implicated in BPDCN pathobiology ^22^. Moreover, potentially oncogenic driver processes affected by these altered profiles included KRAS signaling and the interaction with the extracellular matrix (**Figure 1c**, d).

**Figure 1.**
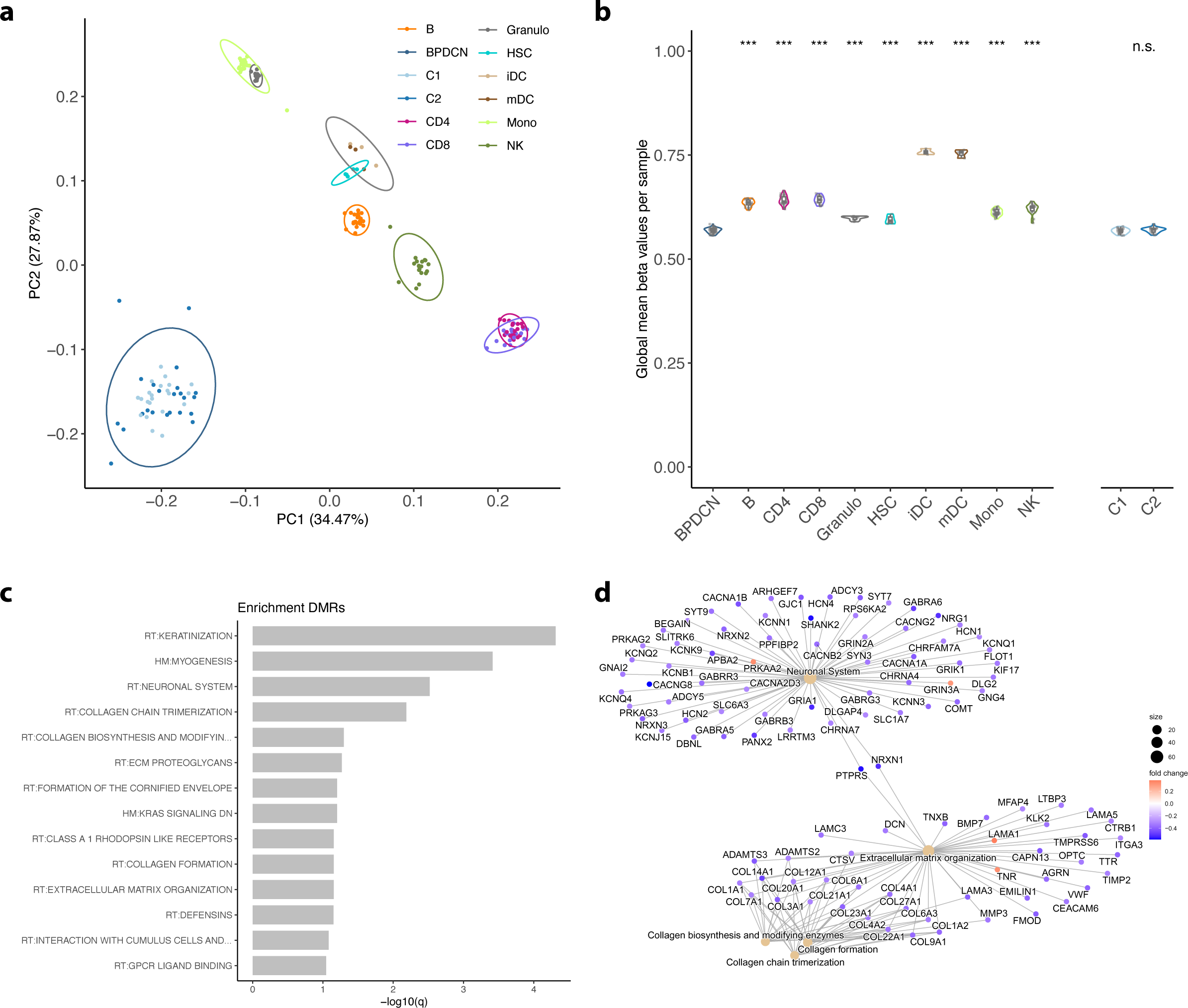
Epigenetic profiles of BPDCN and sorted hematopoietic cell populations. **a** First and second principal components of the 10,000 most variable DNA methylation sites in BPDCN (C1 and C2) and various hematopoietic cell types (B = B lymphocytes, Granulo = granulocytes, HSC = hematopoietic stem cells, iDC = immature dendritic cells, mDC = mature dendritic cells, Mono = monocytes, NK = natural killer cells; ellipses show 95% confidence intervals of multivariate normal distribution). **b** Average genome-wide DNA methylation level (beta values) of BPDCN and various cell types and of BPDCN cluster C1 and C2. Individual estimates are shown as dots and cell type-specific distributions are shown as box- and violin-plots; significant differences against BPDCN were assessed by unpaired Wilcoxon test and significant levels are indicated by asterisks (* = p < 0.05, ** = p < 0.01, and *** = p < 0.001). **c** Enrichment analysis of differentially methylated regions (DMRs) between BPDCN and dendritic cells (harmonic mean of the individual CpG FDRs < 0.01, absolute difference above 0.3) against HALLMARK and REACTOME gene sets. Only significant gene sets are shown (FDR < 0.1). **d** Network enrichment against REACTOME for DMRs (as in **c**); fold changes of DMRs (BPDCN vs DC) are color-scaled (red: higher DNA methylation in BPDCN; blue: higher DNA methylation in DC) and gene sets are denoted by light-brown nodes.

### BPDCN is characterized by a DNA methylation profile distinct from its related entities yet borderline cases exist

To assess the potential of DNA methylation analysis in the differential diagnosis between BPDCN and AML as well as CMML, we performed a comparative analysis of DNA methylation data between our cohort and a previously published cohort of AML patients, representing all relevant molecular subtypes, reanalyzing the BEAT-AML cohort as well as a CMML cohort previously published by Palomo *et al*. This analysis revealed a separation of samples into entity-specific DNA methylation classes in both an unsupervised (PCA; **Figure 2a**) as well as a supervised (PLS-DA; **Figure 2b**) approach. Distinction from T-cell acute lymphoblastic leukemia (T-ALL) and malignant melanoma, which was added for comparison, as another entity originating in the skin, driven by a UV-related mutational signature, was even more pronounced (**Figure 2j**). Intriguingly, we observed four borderline cases through our DNA methylation-based classification (**Figure 2c**; BPDCN_01; 15; 27; 37), exhibiting a relevant amount of overlap between both groups in terms of DNA methylation. In contrast to the overall clear distinction between BPDCN and AML, we found three of these cases to exhibit synchronous concurrent manifestations of AML and/or transformed CMML with pDC features and one of these three cases even presented with genetic features typically encountered in AML with pDC-features (e.g., a *RUNX1* mutation; **Figure 2c**, d-i). The one patient allocated closest to the 95% CI cut-off for the AML definition by our analysis, exhibited a typical pDC-like C1-BPDCN phenotype, while the other, more pronounced borderline classified cases were C2 cases, exhibiting a more immature DC phenotype with fractional monocyte signatures. Subsequent comparative pathway enrichment analysis against HALLMARK and REACTOME gene sets for most differentially methylated regions (DMRs) within gene-body regions revealed a significant enrichment across RHO GTPases, cell migration control and leukemic stem cell maintenance (via HSF1 activation) in BPDCN, whereas promotor regions in BPDCN compared to AML were methylated to a significantly higher degree in epigenetic and transcriptional regulation as well as TP53 regulation and cell cycle control. (**Figure 2m-o**).

**Figure 2.**
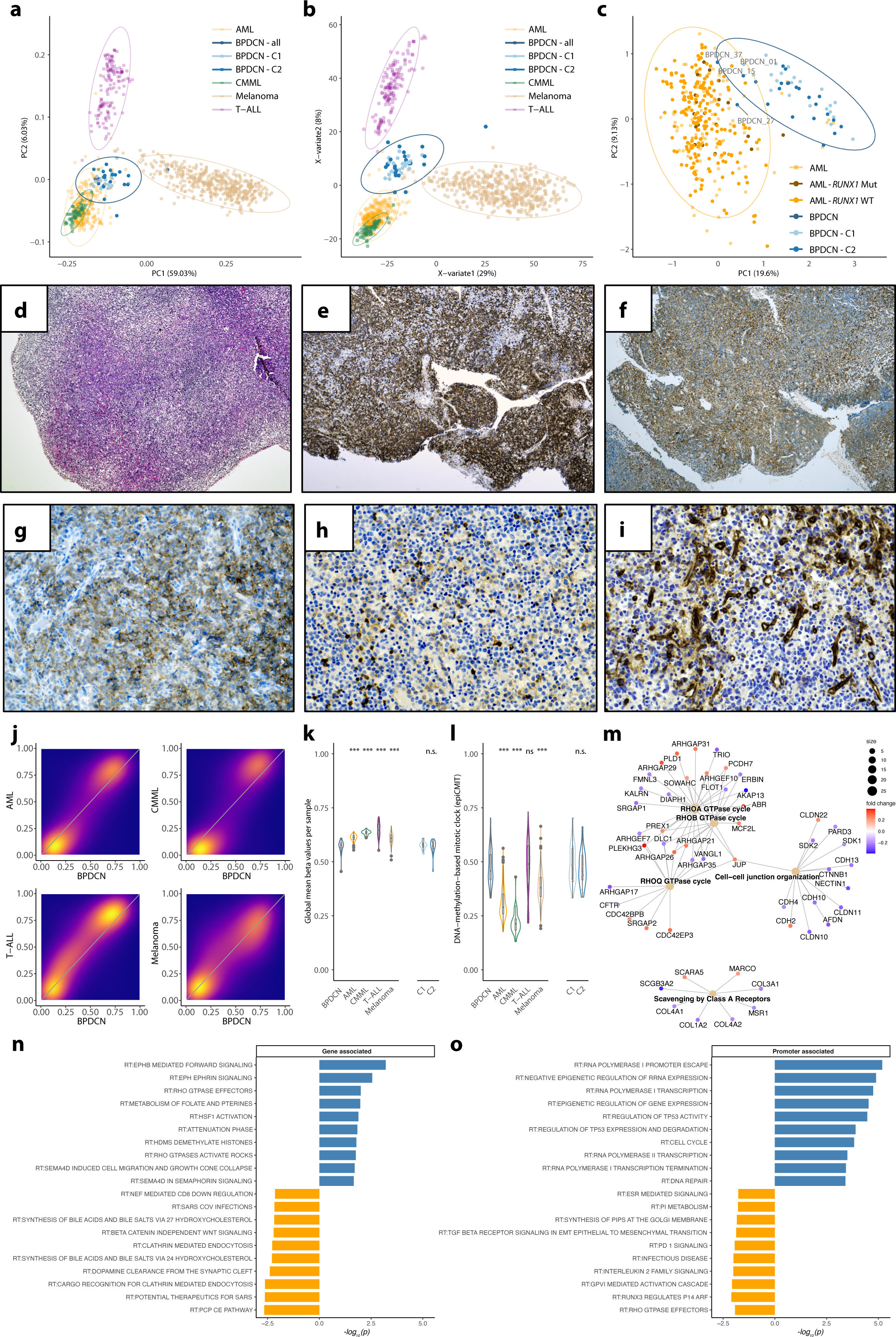
BPDCN DNA methylation in comparison to AML, CMML, t-ALL and melanoma. **a** Visualization of the first and second principal components of the 10,000 most variable DNA methylation sites (ellipses show 95% confidence intervals of multivariate normal distribution). **b** Partial-least squares discriminant analysis (PLS-DA) of adjusted beta values. **c** First and second principal components of the comparison between BPDCN and AML (*RUNX1* wild-type and mutated samples highlighted differently). The four BPDCN cases falling inside the 95% confidence interval of the AML data are labeled. (d-j) a prototypical borderline case with both typical BPDCN as well as AML with pDC-like features; **d** Morphology of the neoplastic infiltrate within the lymph node resembles acute leukemia with polymorphic blast-like cells of variable size (H&E, 40x). **e** Uniform expression of CD123 initially led to the inclusion of BPDCN into the differential diagnosis (CD123, 40x). **f** Further immunophenotypic work-up revealed several atypical features, reminiscent of AML with partial pDC phenotype, including variable expression of CD33 in a significant fraction of the malignant infiltrate (CD33, 40x), yet only partial expression of CD56 (**g**; CD56, 200x) and CD117 (**h**; CD117, 200x). **i** The bi-phenotypic character of the infiltrate is further underlined by a strong CD34 expression of a minor fraction of the blast-like cells alongside the vascular structures, resembling the pDC-like AML phenotype component, whereas the majority of blasts resemble phenotypically characteristic BPDCN cells. **j** Two-dimensional density plots of average CpG site DNA methylation in BPDCN *vs* AML, CMML, t-ALL and melanoma (low density: orchid; high density: yellow/orange). **k** Average genome-wide DNA methylation level (beta values) of BPDCN, AML, CMML, t-ALL and melanoma; for BPDCN subcluster estimates are shown as well. **l** DNA-methylation-based mitotic clock (epiCMIT) estimates for each entity and BPDCN subtypes. **m** Network enrichment against REACTOME for DMRs between BPDCN and AML; fold changes are color-scaled (red: higher DNA methylation in BPDCN; blue: higher DNA methylation in AML) and gene sets are denoted by light-brown nodes. **n** Pathway enrichment against REACTOME gene sets of gene-associated CpGs between BPDCN (blue) and AML (orange). **o** Pathway enrichment against REACTOME gene sets of promotor-associated CpGs between BPDCN (blue) and AML (orange). If not stated differently, differences between BPDCN and the 4 other entities were assessed by unpaired Wilcoxon test and significant levels are indicated by asterisks (* = p < 0.05, ** = p < 0.01, and *** = p < 0.001).

### The BPDCN genome is characterized by an exceptional degree of DNA methylation loss and epigenetic signs of mitotic stress

Global DNA methylation is commonly diminished in malignant compared to healthy cells, being stably maintained in the latter. In this line, BPDCN revealed even the highest methylation loss compared to other blood cancers, including AML, CMML, and T-ALL^23–25^. Global methylation levels in BPDCN even fall below that of melanoma, irrespective of the C1 or C2 subtype (Wilcoxon rank-sum test, p < 0.001). Melanoma was chosen for comparison because of its origin from UV-light exposure in the skin and the ensuing overlap in the mutational signatures (**Figure 2k**)^8, 26^; transcriptional BPDCN subtype (C1 vs C2) did not affect this. Intriguingly, the aggressive clinical nature of BPDCN was further reflected in our observations made through the application of epiCMIT, a DNA methylation-based mitotic clock, which recapitulates the proliferative history of a given tumor sample ^27^. epiCMIT predicted a significantly accelerated mitotic history in BPDCN compared to CMML and even AML, matched only by T-ALL (**Figure 2l**). Although an independent prognostic impact of a high epiCMIT score was observed for a wide range of blood cancers, we observed no such trend in our cohort, plausibly attributable to the limited sample size.

### DNA methylation of tumor suppressor genes is highly deregulated in BPDCN compared to CMML and AML

In keeping with a substantially deregulated DNA methylation profile in BPDCN and a global loss of DNA methylation, we observed promotor regions of tumor suppressor genes to be methylated to an exceptionally high degree, compared to CMML and even AML (**Figure 3a**). At the same time a significantly lower level of gene-body associated methylation in the same genes was identified, again in comparison with CMML and even AML (**Figure 3b**), indicating a substantial oncogenic impact of deregulated DNA methylation in BPDCN (manually selected candidate TSGs, **Figure 3c**, for exhaustive information on deregulated TSGs see **Supplementary Figure 2**). Strikingly, we hereby observe an increasingly deregulated DNA methylation profile from CMML to BPDCN in parallel to the aggressiveness of the entity’s clinical behavior, ranging from the rather indolent and slowly progressing course of CMML in comparison to the more aggressive clinical presentation of AML, which is even more pronounced in BPDCN.

**Figure 3.**
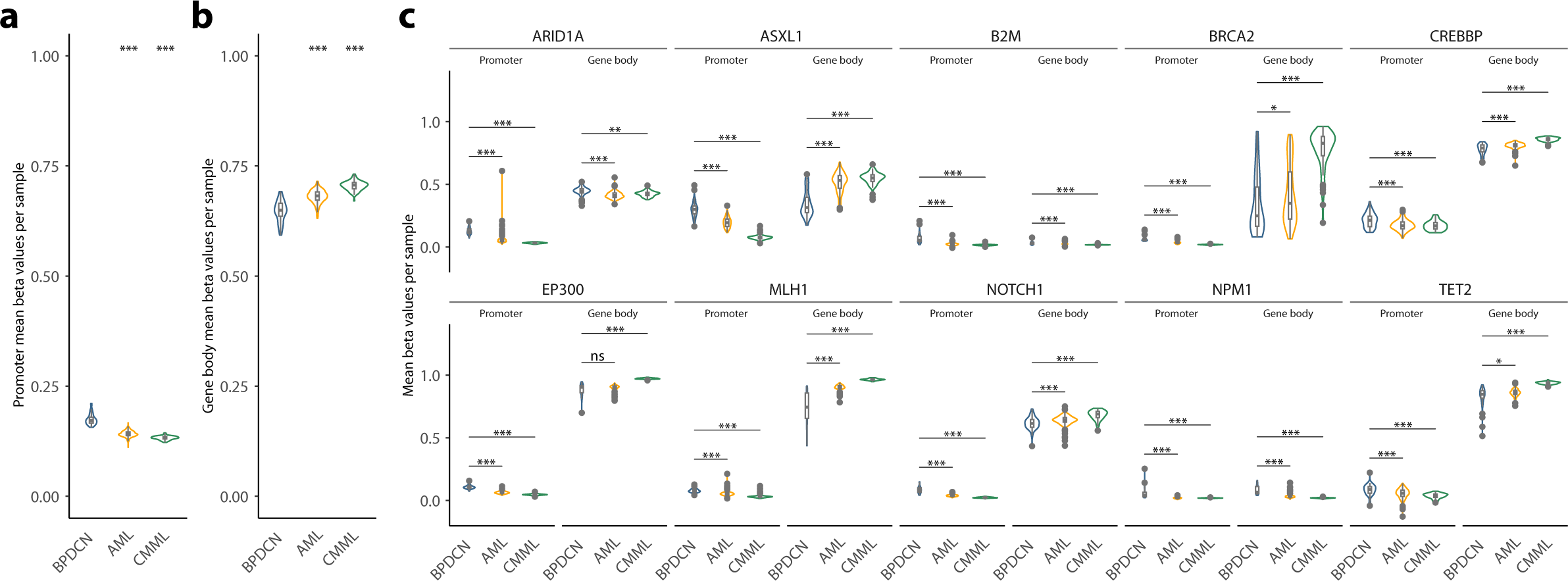
DNA methylation of tumor suppressor genes in BPDCN, AML and CMML. **a** Average promoter DNA methylation (beta values) of tumor suppressor genes in BPDCN, AML and CMML. b Average gene body DNA methylation (beta values) of tumor suppressor genes in BPDCN, AML and CMML. c Average promoter and gene body DNA methylation in selected tumor suppressor genes (for a complete representation of significantly divergent TSGs see **Supplementary Figure 2**). Differences between BPDCN and the AML/CMML were assessed by unpaired Wilcoxon test and significant levels are indicated by asterisks (* = p < 0.05, ** = p < 0.01, and *** = p < 0.001).

### Differential DNA methylation patterns and gene expression signatures differentiate C1 and C2 subtypes in BPDCN and shape a JAK/STAT-driven profile in C2-BPDCN

Building on our previous BPDCN subtype classification, we performed a differential DNA methylation analysis. Alongside differentially mutated genes in BPDCN, we discovered 114 probes, which were differentially methylated between C1 and C2 (p < 0.0001; 10,136 probes with p < 0.01), corresponding to a relatively similar methylome, in keeping with our above PCA/PLS-DA (depicted as scales beta values in **Figure 4a**; beta values see **Supplementary Table 4**).

**Figure 4.**
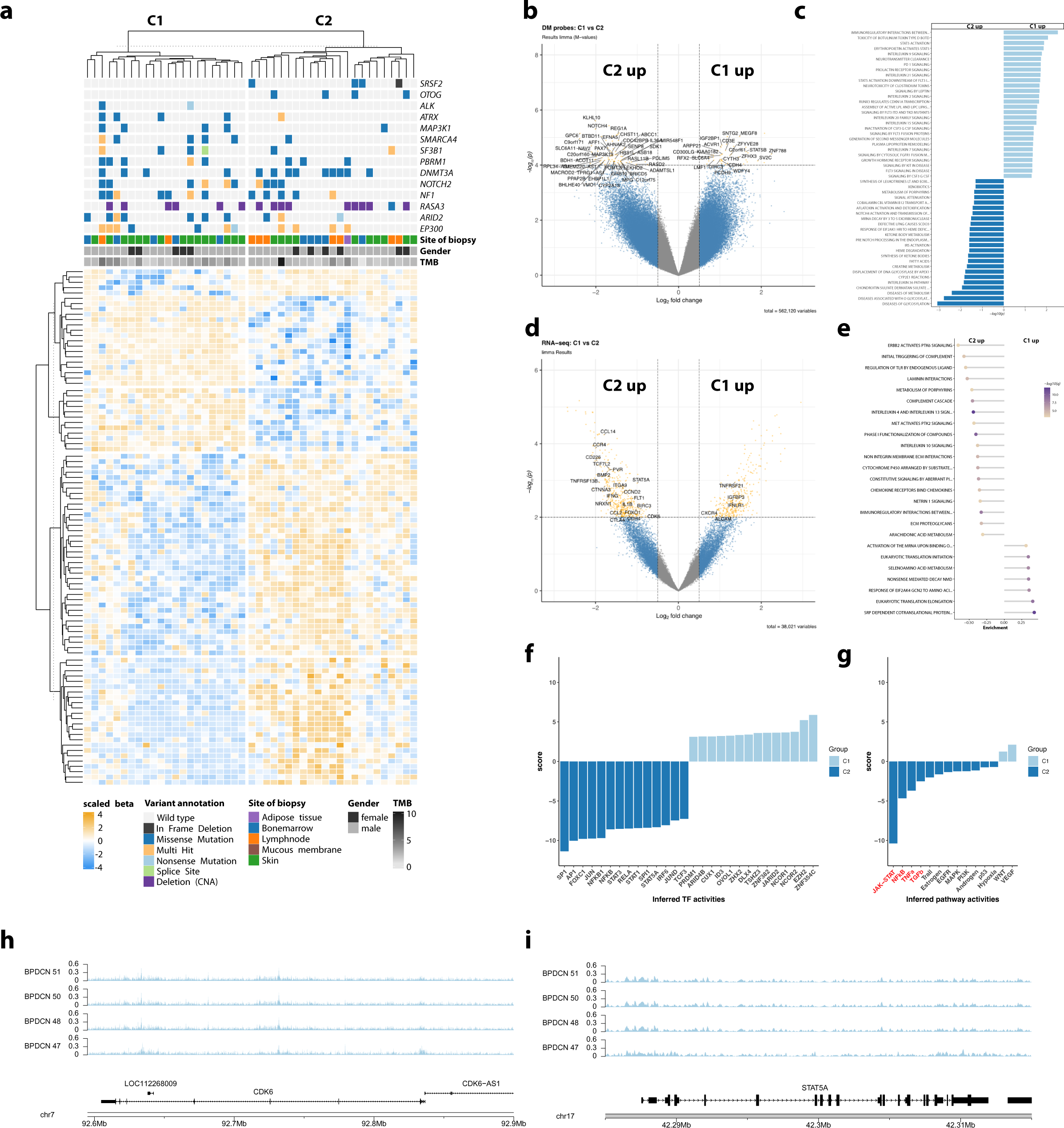
Differential DNA methylation, expression and mutation patterns in BPDCN subtypes C1 and C2. **a** Top part of the heatmap shows mutational patterns of 14 genes previously identified as significantly enriched between the two clusters ^6^. The bottom part shows scaled beta values (DNA methylation levels) of differentially methylated CpG sites (p < 0.0001). **b** Volcano plot of beta values showing log_2_ fold-changes and p-values with gene annotations for significantly different CpGs (p < 0.0001). **c** Gene set enrichment analysis results of DNA methylation data against REACTOME gene sets (p < 0.05). **d** Volcano plot of expression profiles showing log_2_ fold-changes and p-values with gene annotations for significant differentially expressed tumor suppressors genes, oncogenes and genes involved in cell adhesion/cell cycle (p < 0.0001). **e** Gene set enrichment analysis of RNA-seq data against REACTOME gene sets (p_adj_ < 0.001, absolute enrichment > 0.3). **f** Transcription factors with significantly different inferred activity (p < 0.05) in C1 (light blue) and C2 (dark blue). **g** Pathway activities in C1 (light blue) and C2 (dark blue) with significantly different pathway activities shown in red font (p < 0.05). **h,i** FFPE-ATAC-seq estimates of chromatin accessibility for *CDK6* and *STAT5A* for four BPDCN cases belonging to subcluster C1. CDK6 is located on the minus strand and *STAT5B* on the plus strand, respectively.

Subsequently, to gain insight into the effect of differential DNA methylation on the phenotypical development of both subtypes, we assigned differentially methylated probes to their respective genes (**Figure 4b**). Gene set testing (GST) (**Figure 4c**) revealed enrichment in elevated DNA methylation levels for interleukin signaling genes as well as prominent members of JAK/STAT signaling (including STAT5B) in C1-BPDCN, whereas C2-BPDCN samples exhibit significantly pronounced DNA methylation of posttranslational modifications like glycosylation and metabolic processes (e.g., vitamin and heme metabolism).

Expanding on our differential assessment of C1 and C2-BPDCN onto the transcriptional level, we performed differential gene expression analysis (**Figure 4d**, e). Hereby, we observed an induction of innate and adaptive immunological processes alongside an upregulation of extracellular matrix-interactions in C2 cases by gene set enrichment. Further, we found several prominent candidate genes, including *STAT5A*, *CDK6*, *CCR4*, *CCND2* and *FOXO1* to be expressed at significantly higher levels in atypical C2 cases. Corresponding to higher gene expressions in C2, *CDK6* (p = 5.3×10^-4^, log_2_ fold-change 1.52) and *STAT5A* (p = 5.5×10^-3^, log_2_ fold-change 1.19) had promotor-associated sites that were significantly higher methylated in C1-BPDCN leading to relative transcriptional inactivation in C1. Correlation of DNA methylation and relative RNA-seq derived gene expression data yielded a substantial number of significant correlations among TSGs and oncogenes including *BCL2*, *STAT5A* and *SOX1* (**Supplementary Table 5a**). Upon extension of the analysis unto the entire transcriptome further significant correlations (q < 0.1) were observed (**Supplementary Table 5b**).

In order to further focus our observations on potential therapeutic applicability, we inferred transcription factor (TF) activities from bulk RNA-seq data employing CollecTRI as a resource for TFi pathway activity inference with PROGENy. Hereby we observed the transcriptional mirror image of the divergence in DNA methylation profiles between C1 and C2-BPDCN. In particular, we observed significantly higher activity of NFkB (driven by FOXC1, NFKB1 and NFKB; p < 0.05) and more strikingly JAK-STAT (predominantly driven by STAT3, STAT1 and STAT5A; p < 0.05) associated TF in C2 and an EZH2 dependence in C1-BPDCN (**Figure 4f**, g). In addition, we supplemented these observations by FFPE-ATAC-seq of four C2-BPDCN, which revealed substantial chromatin accessibility, in keeping with our epigenetic and transcriptional findings (**Figure 4h**, i).

### Cellular composition of the tumor microenvironment by MethylCIBERSORT and immunohistochemistry reveals distinct immunological subtypes correlated with tumor genomics

In order to evaluate the composition of the tumor microenvironment (TME), we conducted a MethylCIBERSORT analysis on all 45 high-quality genome-wide DNA methylation profiles from primary diagnostic samples. From the inferred relative abundance of T cells and their respective subpopulations (CD4^+^ effector cells, CD8^+^ cytotoxic cells, regulatory T cells (Tregs)) alongside B-cells, natural killer (NK)-cells neutrophils, monocytes, eosinophils and stromal cells (fibroblasts, endothelial cells) two TME classes (IC1 and IC2) were predicted. The smaller class (IC1) exhibited a depletion in monocytes, B- and NK-cells, an enrichment in Tregs and a trend towards higher counts of neutrophils and cytotoxic T cells (**Figure 5a**). These observations were subsequently validated via IHC for tumor-infiltrating T cells and monocytes with a significant correlation between relative distributions of cell populations according to the respective methodology (**Figure 5b** – e, g, h). Comparing IHC and MethylCibersort estimates, we found a significant correlation in monocytes (Pearson correlation coefficient = 0.6437, p = 1.84×10^-6^) and in T-cell populations (Pearson correlation coefficient = 0.3809 p = 0.0098). Beyond a statistical trend towards an overall higher TMB, we observed an enrichment in mutations affecting *ERBB2*, *ASXL1, EP300* and *KMT2C* mutations. Further, we identified more frequent mutations in *CDH1, JAK2, SMAD2, NOTCH1* and *DNMT3A* (**Figure 5** f). Patients in the IC1 subgroup had a significantly shorter progression-free survival (p = 0.039) and a trend towards inferior overall survival (p = 0.14) in IC1 patients (**Figure 5** i, j). Correlating the results from our MethylCIBERSORT approach with the genomic landscape of BPDCN (restricted to on genes mutated in at least 30% of cases; n = 8), mutations in *EP300*, *KMT2C* and *NOTCH2* were associated with a significant enrichment of eosinophils (*KMT2C*: p = 0.038), monocytes (*EP300*: p = 0.045; *NOTCH2*: p =0.0026), CD4 effector cells (*EP300*: p = 0.042) and regulatory T-cells (*EP300*: p = 0.012) within the TME, respectively (**Supplementary Figure 4**). Previous reports identified a strong correlation between DNA methylation and chronological age ^28, 29^. Based on our MethylCIBERSORT results leading to the allocation of two clusters, epiCMIT was applied to estimate the history of proliferative stress/DNAm age. In contrast to the genome/transcriptome-based BPDCN clusters C1/C2 (**Figure 2l**), the immunological cluster IC2 showed a DNA methylation imprint of significantly higher proliferative stress compared to IC1, which resembles DNA methylation-based pre-aging in this subgroup (p = 0.038; **Supplementary Figure 5**).

**Figure 5.**
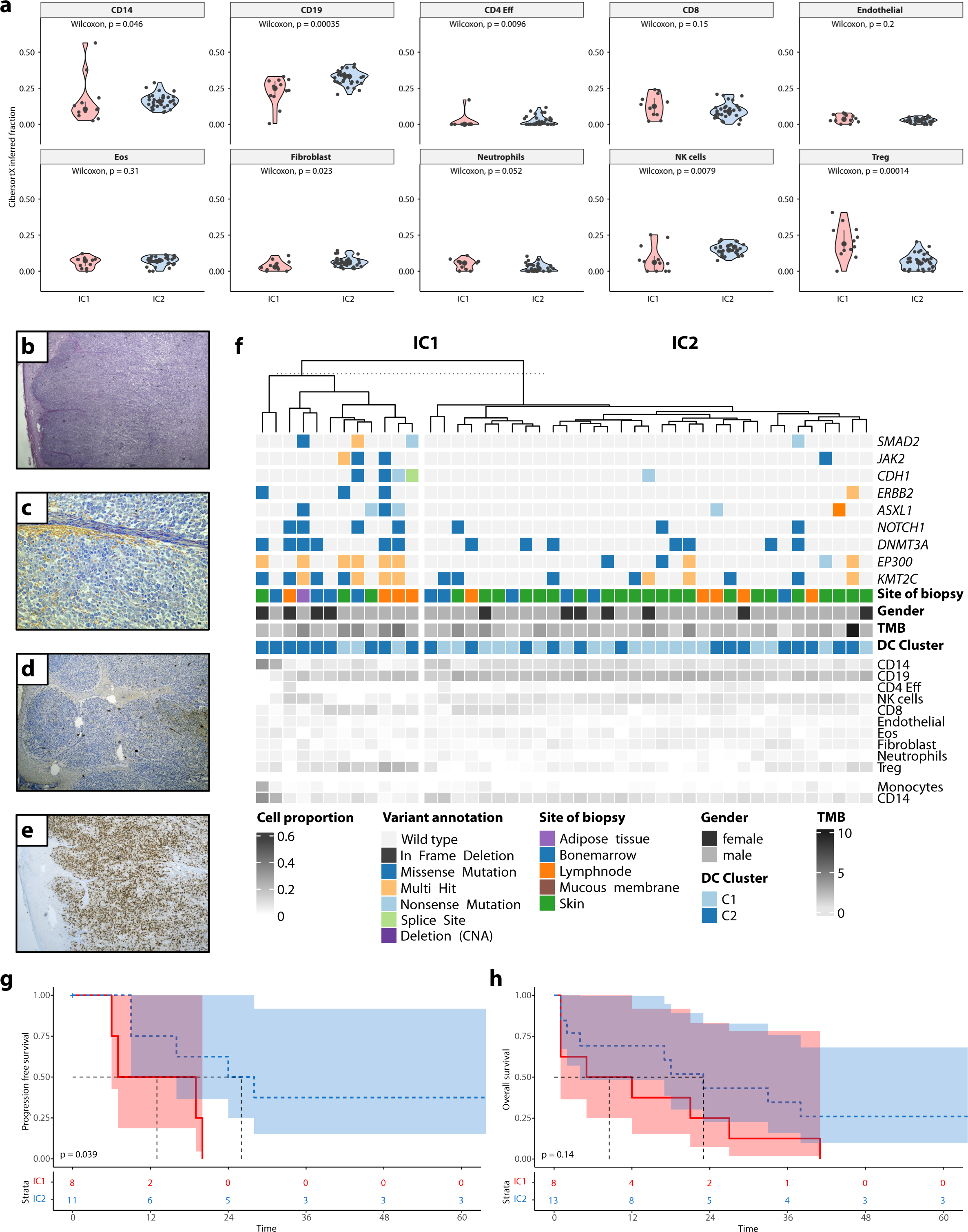
Tumor immune composition by MethylCIBERSORT identifies clusters of divergent immunogenicity. **a** DNA methylation data were deconvoluted according to immune cell populations (MethylCIBERSORT). This revealed two different types of BPDCN (Treg and CD14 driven; named IC1 and IC2, respectively) presenting with significantly differing immune cell subsets in regard to the markers CD14 (monocytes/macrophages), CD19 (B-cells), CD4 (T-helper cells) as well as fibroblasts, NK-cells and T-regulatory cells (T-regs). **b – e** A borderline BPDCN/AML pDC-like case analyzed by MethylCIBERSORT and a comparative immunohistochemical assessment of the tumor microenvironment is presented. **b** H&E staining reveals a cutaneous infiltrate covered by an intact epidermis. **c** Giemsa staining reveals small blastoid cells with partly roundish occasionally monocytoid nuclei, small nucleoli and weakly basophilic cytoplasm with increased mitotic activity. **d** Staining for myeloid peroxidase reveals expected negativity in the malignant infiltrate alongside a few positive, tumor-infiltrating myeloid cells. **e** However, CD14-Expression highlights both a typical negative BPDCN population, as well as a relevant monocytoid population, including few tumor-infiltrating monocytes alongside a larger subgroup of malignant cells. **f** Oncoplot displays mutational patterns of 9 genes that were found to be more differentially mutated between IC1 and IC2. Additionally, the heatmap illustrates TME cell proportions for each individual sample. **g, h** Progression-free (PFS) and Overall survival (OS) analysis for patients with available clinical follow-up according to IC1 vs IC2 identifies a significant inferior prognostic impact for the IC1 subtype regarding PFS accompanied by a trend towards inferior OS.

In keeping with the higher levels of DNA methylation and the subsequently suppressed expression of adaptive immunological processes in typical pDC-like C1-BPDCN cases, we observed a trend towards higher levels of tumor-infiltrating T-cells in C2-BPDCN samples despite the overall higher TMB in C1 patients, regardless of patient age or location of the tumor sample by both MethylCIBERSORT and IHC (**Supplementary Figure 6a, b**).

### DNA methylation-based clusters cannot be fully recapitulated in transcriptional BPDCN subtypes

The most 5,600 variable CpG probes were subject to discovering DNA methylation subtypes using K-means clustering. The optimal number of clusters was determined using the average silhouette method and gap statistics. Both methods agreed on the optimal number of clusters (n = 2). Upon unsupervised cluster analysis, we observed no statistically significant recapitulation of transcriptional BPDCN subtypes C1 and C2 in the methylome. Some overlap was, however, found in shared genomic features (*EP300* and *ATRX* mutations in C1 and MethC1), consistent with higher TMB in C1 and MethC1. A significantly more pronounced overlap was apparent upon correlation analysis between DNA methylation-based cluster allocation and immunological clusters according to our methyCIBERSORT-derived immunoclusters (**Supplementary Figure 7a,b**).

### Mutational drivers and promotor status of epigenetic regulators shape the proliferative fate and DNA methylation profile of BPDCN

In order to provide a genomic context for the DNA methylation profiles obtained within this study, we performed WES on all patients, who were not part of our previous molecular landscape project in BPDCN, which led to the most comprehensive mutational characterization of any BPDCN cohort to date (**Figure 6a**). WES of our extended BPDCN cohort identified a higher mutational load compared to the C2 cluster. This further supports the assumption of another non-mutational mechanism driving in C2-BPDCN such as a deregulated DNA methylation and transcriptional profile outlined above. The exceptional pathophysiological role of epigenetic features within the C2 cluster is supported by more frequent *DNMT3A* alterations bordering on statistical significance in this limited cohort and the significant deregulation of splicing genes such as *SRSF2*.

**Figure 6.**
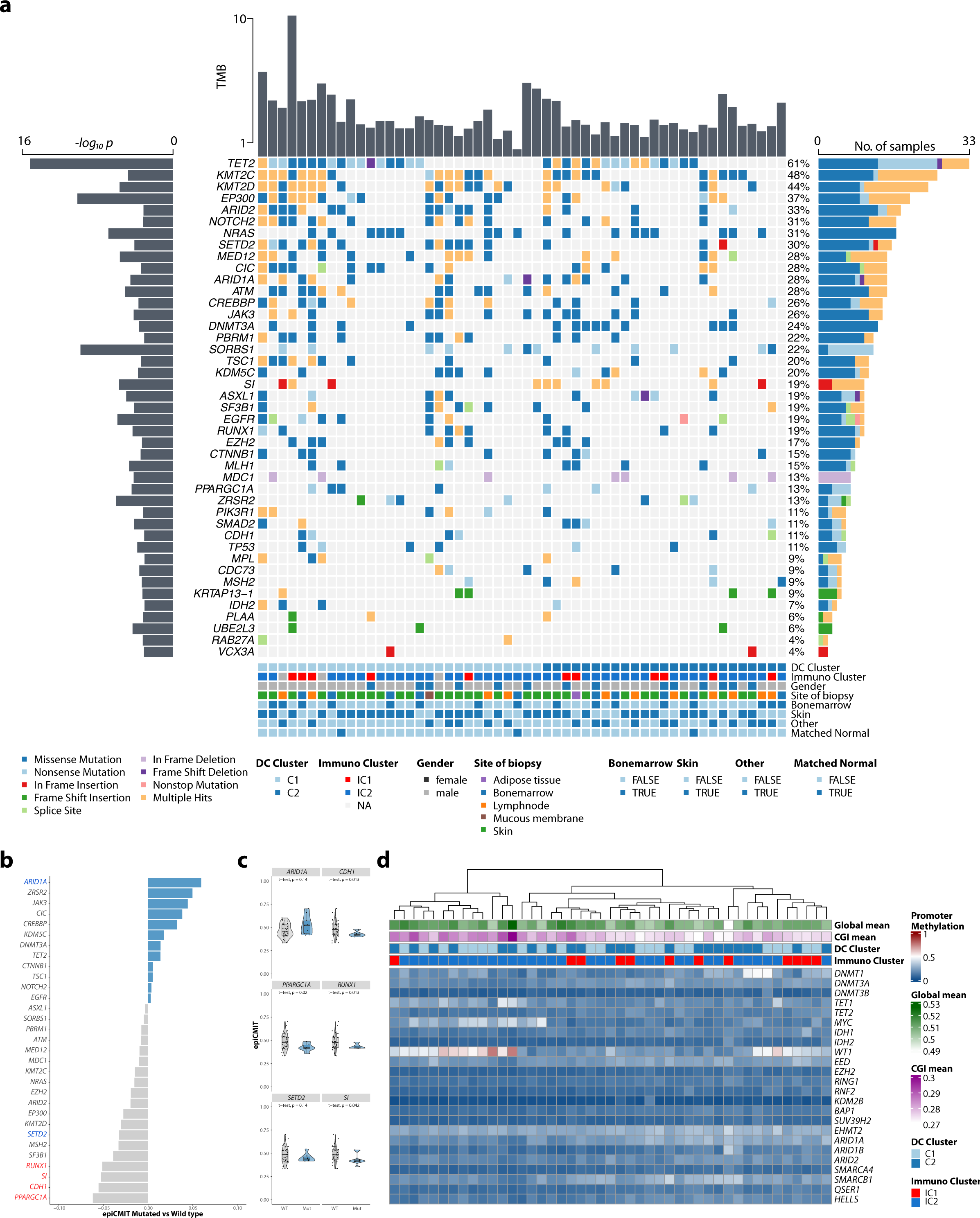
Impact of significant driver mutations and DNA methylation status of epigenetic regulators on proliferative history and global CGI DNA methylation. (**a**) Oncoplot displaying putative driver genes inferred by MUTSIGCV analysis in addition to tumor mutational burden (TMB; upper bar plot), *-log_10_* p-values (left bars) and the number of samples harboring mutations in a given gene (right bar). In total **6.065** bioinformatically deleterious mutations, in **3,970** genes were observed. Of these indels and SNVs, 4,999 were missense (82.4%), 321 nonsense (5.3%), 12 non-stop (0.2%) and 487 indel mutations (8.0%). Mutation types are color-coded, and covariates are shown below for each sample (covariate ‘Other’ refers to samples with tissue affected other than skin or bone marrow). (**b**) Analysis of which significant mutational driver alterations may confer a higher or lower proliferative capacity to BPDCN cells. Statistically significant associations are delineated in red and blue, respectively. (**c**) Comparative analysis regarding proliferative capacity in selected genes’ mutant and wild-type status. (**d**) A heatmap depicting the DNA methylation status of gene promoter regions across a set of preselected epigenetic regulators associated directly and indirectly with global DNA methylation, as described ^45^. Average global DNA methylation and CpG islands (CGI) are estimated per patient.

Following mutational characterization, we wondered whether specific mutational drivers were associated with the proliferative history of a given case, thereby integrating mutational and epigenetic datasets in order to identify genetic alterations, which could confer a selective advantage as mitotic accelerators in BPDCN cells. While definitive conclusions are complicated by the limited sample size, we observed a trend towards a proliferative increase in cases harboring *ARID1A*, *ZRSR2*, *JAK3*, *CIC* and *CREBBP* mutations and a (significant) decrease in mitotic activity in cases affected by mutations in *PPARGC1A*, *CDH1*, *SI*, *RUNX1*, *SF3B1*, *MSH2* and *SETD2* (**Figure 6b**, c). In summary, our findings suggest, that the proliferative potential of BPDCN is determined by certain mutational driver events.

In order to unravel additional determinants, which shape the overall strongly demethylated landscape in BPDCN, we investigated the promotor DNA methylation status of a set of genes associated with epigenetic regulation (**Figure 6d**). Hypermethylation of the promotor region (promoter CGI methylation >0.2) of *DNMT1*, *WT1*, *MYC* and *TET1* was hereby observed in 42.2%, 73.2%, 40.0% and 53.3% of patients respectively. Further, we observed a significant correlation between promotor methylation and overall CGI methylation for 22 of the 24 genes (Spearman’s rank correlation, p_adj_ < 0.05) signifying a substantial impact on the epigenetic landscape in BPDCN. Only DNMT1 and KMD2B did not show a significant correlation. Of note, a substantial overlap between patients harboring *WT1* and *MYC* promoter methylation was observed 16/45 cases (35.6%). Given that *MYC* constitutes a known *WT1* target, this hints at a coupled mechanism of both genes in BPDCN pathogenesis ^30^ (**Figure 6b, c**).

## Discussion

BPDCN is a clinically and molecularly heterogeneous disease that poses a multi-level challenge in terms of diagnostics and treatment alike. Unraveling the epigenetic characteristics of the disease may pose an aid in providing a correct and timely diagnosis, which is crucial for the initiation of specific treatments. Through a comparative analysis of the DNA methylation landscape of BPDCN incorporating related hematological malignancies (AML, CMML and T-ALL patients, encompassing all relevant genetic and phenotypical subgroups), we identify distinguishing features and provide unprecedented insights into the molecular pathogenesis of BPDCN, which may in term facilitate the development of more refined targeted therapeutic approaches. In this integrated molecular study of BPDCN we report on the largest cohort studied so far. Where previous reports on BPDCN were limited by small cohorts and restricted methodological approaches, we systematically defined genomic and transcriptional signatures in the context of genome-wide DNA methylation ^4, 8, 10, 31–37^. Hereby, we made three novel and essential observations.

First, our study of DNA methylation profiles allowed for a clear distinction between globally demethylated BPDCN patient samples with only localized DNA hypermethylation showing dominant signs of mitotic stress, resembling a pronounced variant of the canonical cancer methylome on the one hand and AML, CMML as well as T-ALL on the other. In keeping with previous assumptions, we find enrichment in cases with distinct DNA hypomethylation, predominantly affecting late-replicating regions and molecular signs of extensive past mitotic stress ^38^. Several patients, for whom a borderline DNA methylation profile between AML and BPDCN was identified, were found to resemble cases with either syn- or metachronous development of CMML/AML or a molecular and phenotypical constellation resembling AML with pDC-like features ^9, 10^. Moreover, we observed all but one of these cases to exhibit the atypical, more immature cDC-impacted C2-BPDCN transcriptional profile ^6^. This aligns with observations in B-cell malignancies and solid tumors where the DNA methylation profile reflects the degree of maturation of the cell-of-origin ^27, 39^. Recently, clonal hematopoiesis of indeterminant potential (CHiP), disrupting epigenetic regulators in the majority of BPDCN cases, was proposed as an underlying mechanism rendering secondary mutations in RAS signaling (*NRAS*, *KRAS*) and tumor suppressors like *TP53* and *ATM* secondary clonal events and thereby more specific in BPDCN pathogenesis ^40^. Our previous genomic studies revealed a significant overlap in mutational drivers between BPDCN and the abovementioned entities, underscoring the close molecular relatedness between BPDCN and especially AML/CMML, which was recently further illustrated in cases of divergent clonal evolution from a CHiP constellation. The present study of DNA methylation signatures, however, revealed a significant impact on signaling processes ultimately contributing to the specific phenotype of

BPDCN in particular. This can therefore be harnessed as a reliable discriminator beyond established immunophenotypical and histopathological approaches ^23, 41, 42^. Moreover, DNA methylation profiling revealed a canonical epigenetic deregulation of TSGs with putative oncogenic effects, vastly exceeding previous observations in related entities ^23, 24^. Intriguingly, we observed several mutational drivers of mitotic stress, signified through a proliferative epigenetic signature. In line with previously reported effects of *ARID1A* mutations on cell proliferation, we found *ARID1A* mutant BPDCN samples to exhibit pronounced signs of proliferative activity, while decreased mitotic activity in *RUNX1* mutant BPDCN is in keeping with reduced proliferation in hematopoietic stem cells harboring similar mutations ^43, 44^. At the same time, unsupervised clustering of genome-wide DNA methylation levels only partially recapitulated transcriptional clusters, which was, however, previously observed in other blood cancers (e.g., T-ALL) as well ^45^.

Second, our recently established molecular subgroups of BPDCN differ in terms of both transcriptional profile by RNA-seq and to a lesser extent by DNA methylation signature. Confirming and expanding on previous studies, we found predominant pathways deregulated by these circumstances to include both innate and adaptive immunological processes, for which gene expression was significantly induced in C2-BPDCN cases. This corresponds well to elevated DNA methylation levels in interleukin/inflammatory signaling genes in C1-BPDCN, leading to a relative up-regulation of the interleukin 4/13 interactome in C2-BPDCN ^46^. Interrogating our bulk RNA-seq data for TF activity, we observed the transcriptional mirror image of the divergence in DNA methylation profiles between C1 and C2-BPDCN. Of particular interest, regarding potential therapeutically targetable vulnerabilities, we observed significantly higher activity of NFkB (driven by *FOXC1*, *NFKB1* and *NFKB*) and more strikingly, JAK-STAT (predominantly driven by *STAT3*, *STAT1* and *STAT5A*) associated TF activity in C2 in contrast to an *EZH2* dependence in C1-BPDCN. Potent and clinically well-tested inhibitors for potential molecularly informed therapeutic combinations with tagraxofusp are readily available ^47^. These observations were then verified by FFPE-ATAC-seq, which revealed substantial chromatin accessibility at highly expressed loci, including *CDK6* and *STAT5A*, in keeping with our epigenetic and transcriptional findings.

Finally, we gained insight into the tumor immune microenvironment through the combined analysis of MethylCIBERSORT and immunohistochemistry. The essential finding was that there are two, prognostically relevant immunologic clusters (IC1 and IC2) in BPDCN, characterizing the unfavorable IC1 subgroup (comprising approx. 25% of patients) harboring a trend towards a higher TMB and significant enrichment for *ERBB2*, *ASXL1, EP300* and *KMT2C* mutations alongside a TME relatively depleted of NK-cells, monocytes and B-cells but enriched in Tregs. A previous report raised the issue of immunotherapeutic approaches in BPDCN immunohistochemically investigating PD-L1 expression levels^48^. Our comprehensive MethylCIBERSORT approach identified a relevant subset of immunologic hot cases in which immunotherapeutic strategies seem to be promising in light of lacking treatment options beyond tagraxofusp^48^. Moreover, we found that IC2 had a significantly higher epigentic age but there was no difference regarding chronological age between both subgroups. This is congruent with previous observations in NSCLC^19^, where it was hypothesized that a higher epigentic age was able to emulate the malignant potential of tumors with a high mutational load or decisive driver mutations such as *TP53*. However, the authors went on to demonstrate that a higher epigentic age was associated with favorable survival compared to tumors characterized by genomic instability and high mutational loads^19^. In addition to the immunologically defined clusters, we observed an elevated level of TILs in atypical C2-BPDCN. Our future goal is now to dissect the TME in even greater detail by single-cell RNA sequencing and spatial transcriptomics.

Limitations of the current study include a restricted number of cases alongside a partial lack of information on clinical follow-up for about half of the cohort. Additionally, a substantial subset of tagraxofusp-treated patients will make a valuable addition to future studies.

In conclusion, employing an integrated molecular approach, we are able to reliably distinguish BPDCN from its related entities and identify clinical and molecular borderline cases. Further, we unravel the epigenetic and transcriptional underpinnings of our two recently defined subtypes of BPDCN, identifying divergent potential targetable vulnerabilities and characterizing immunologically and prognostically meaningful subtypes through DNA methylation profiling-derived quantitative TME assessment validated by immunohistochemistry.

## Methods

### Case selection, clinicopathological assessment, whole exome, and whole transcriptome sequencing

For this retrospective analysis, we reviewed our institutional archive for cases of histologically confirmed BPDCN between January 2001 and April 2022. The study was approved by the ethics committee of the University of Lübeck (reference-no 18-311) and conducted in accordance with the declaration of Helsinki. Patients at the Consultation Center for Hematopathology provided written informed consent regarding routine diagnostic and academic assessment, including genomic studies. Histopathological work-up was performed as described^6^ and yielded 74 cases of BPDCN, 54 of which had sufficient FFPE tissue samples available for subsequent molecular analysis were selected and subjected to a comprehensive immunophenotypic workup (47 samples were included in a previously published investigation of the mutational landscape in BPDCN^6^). Genomic DNA and RNA were extracted from three 5µm FFPE tissue sections of either tumor or normal tissue (where available; n = 3) employing Maxwell® RSC DNA FFPE kit and Maxwell® RSC RNA FFPE kit (both Promega). WES and RNA-Seq following library preparation using Agilent SureSelect Human All Exon V6 library preparation kit (Agilent Technologies) and NEBNext® UltraT Directional RNA Library Prep Kit (New England BioLabs), respectively were performed on a NovaSeq platform (Illumina) at Novogene (UK) Co. as described ^49^. Tumor whole exome libraries were sequenced to a median depth of 131x (mean 134 ± 52 s.d.) and normal libraries reached a median depth of 67x (mean 83 ± 36 s.d.).

### Whole exome data processing and variant calling

Sequencing data from WES was processed using the same workflow as previously described^6^. Briefly, raw reads (paired-end fastq format) were trimmed (adapter and quality values) applying FASTP (v0.23.0; minimum length, 50 bp; maximum unqualified bases, 30%; trim tail set to 1)^50^; trimmed reads were mapped to GRCh38 using bwa mem (v0.7.15)^51^ and mappings were converted into BAM format using PICARD TOOLS (v2.18.4). Next, mate-pair information was fixed, PCR duplicates were removed, and base quality recalibration was performed using PICARD TOOLS, GATK (v4.2.3.0)^52^ and dbSNP v138 ^53^. Single nucleotide variants (SNVs) and short insertions and deletions (indels) were identified following GATKs best practices for somatic mutation calling (matched normal-tumor mode for samples with normal tissue available (n=3) and tumor-only mode for samples without normal tissue available). MUTECT2 (GATK)^54^ was applied to the processed mappings with GNOMAD variants as germline resource and the b38 exome panel from the 1000 genome project as a panel of normal, capturing the expected repertoire of germline variants to be expected in a Central European study population. Next, FFPE read orientation artifacts were identified and removed according to GATK guidelines. Filtered variants were annotated using VARIANT EFFECT PREDICTOR^55^ (VEP v103, GRCh38; adding CADD v1.6, dbNSFP v4.1a23, and GNOMAD r3.0 as additional resources) and annotations were converted into *MAF* format using VCF2MAF (V1.6.21) (DOI:10.5281/ZENODO.593251); coverage was extracted directly from the vcf INFO field. The top 20 frequently mutated genes (FLAGS)^56^ were removed from further analysis and the remaining somatic variants were filtered as follows: minimum coverage of 40, minimum alternative allele coverage of 5, minimum variant allele frequency of 10%, and only variants with a frequency < 0.1% in 1000 genomes, GNOMAD, or ExAC were considered for subsequent downstream analysis. High-impact variants (CADD score > 10) in tumor suppressors according to Vogelstein et al. ^57^ were filtered as such that minimum coverage of 20 minimum, minimum alternative coverage, and minimum variant allele frequency of 10% was required. Genes mutated more often than expected were identified by applying MUTSIGCV (v1.41)^58^ and potential drivers were identified using p < 0.001. Tumor mutational burden (TMB) did not differ between tumor-only samples and samples with matched normal tissue at the finalization of the filtering process (Wilcox test, p = 0.1004).

### Transcriptome Data Processing, Quantification, Deconvolution, and Analysis

Gene expression sequencing data were processed in the same manner as described previously^6^. Briefly, gene expression profiles were retrieved from adapter trimmed reads (FASTP as described above) using STAR ALIGNER (v2.7.4b)^59^ against GRCh38 (GENCODE v37) as reference. On average 73.5 million reads (median 81.1) were successfully mapped to the human reference per tumor sample and 29.6 million reads (median 30.4) per normal sample, respectively. Count profiles were normalized applying MIXNORM (v0.0.0.9000; 50 iterations, tolerance set to 0.1) ^60^, which removes unwanted biological and technical effects from FFPE material that can bias the signal of interest. Differentially expressed genes on normalized expression values between two conditions were identified using a linear modeling approach (LIMMA package, v3.50.1)^61^.

Pathway enrichment analysis against REACTOME gene sets (MSIGDF R package v7.4) on significantly differentially expressed genes was performed using a rank-MANOVA-based approach as implemented in MITCH (v1.12.0; priority on significance) ^62^; NF-Kβ pathway (KEGG) was added manually to the REACTOME gene set.

Previously published scRNA-data from Villani et al.^63^ was used to infer the cell-type composition of the bulk RNA-Seq data with respect to dendritic cells and monocytes applying a deconvolution of damped weighted least squares (DWLS) method as described previously ^6^.

### Transcription factor and Pathway activities from RNA-seq data

Transcription factor activities were inferred from RNA-seq data using prior knowledge as provided by Collection of Transcriptional Regulatory Interactions (CollecTRI (Preprint at https://doi.org/10.1101/2023.03.30.534849 (2023)), DECOUPLER v2.6.0), which provide a curated collection of 1,175 transcription factors. Briefly, t-values between two conditions (BPDCN DC cluster) were estimated using LIMMA. Transcription factor activities were estimated on *t*-values using weighted means and activities with p < 0.05 were plotted (28 transcription factors).

Pathway activities for curated pathways with weights for each interaction were estimated using Pathway RespOnsive GENes for activity inference (PROGENy ^64^, gene signatures for 14 pathways) on limma estimated *t*-values (as above).

### Genome-wide DNA methylation profiling and data analysis

Whole-genome DNA methylation analysis was carried out on all 54 cases of the study cohort employing the Illumina EPIC array at ATLAS Biolabs. Bioconductor R package MINIFI (v1.46.0) was used to further process raw IDATs that were previously generated from iScan. The quality of samples was checked by using mean detection *P*-values and only samples with *P*-values <0.05 were kept for further processing (five samples excluded). In addition, according to in-house bioinformatic QC pipelines (Glaser *et al.*, in preparation) was applied (one additional case excluded). Remaining samples were normalized using quantile normalization (funtion *preprocessQuantile*) and DNA methylation data predicted sex was compared to the actual sex. Samples, where the predicted sex did not match with the actual sex, were removed (four samples removed), leaving 45 samples for further analysis. DNA methylation probes were quality filtered and probes with non-significant *P*-values were removed (*P* > 0.01). Additionally, cross-reactive probes and BOWTIE2 multi-mapped probes were removed, and M- and beta-values of the remaining probes were extracted ^65^.

Differentially methylated probes between two conditions were identified using a linear modeling approach as implemented in LIMMA. Generalized gene set testing on differentially methylated probes was performed by applying the *gsameth* function (MISSMETHYL package v1.34.0) against the REACTOME and/or HALLMARK gene sets (MSigDB v7.5).

### Comparative analysis with genome-wide DNA methylation data from different cell types

To assess epigenetic differences between BPDCN and several cell types, we used data from different sources. Raw DNA methylation data (IDAT files) from B-lymphoid cells (n = 23), naïve CD4+ (n = 23), naïve CD8+ cells (n = 19), granulocytes (n = 10), monocytes (n = 24) and NK-cells (n = 20) was retrieved from GEO accession GSE184269 ^20^. Data were processed as described above with the exception that normalization was performed using subset-quantile within array normalization (*SWAN*, MISSMETHYL) and beta-values were extracted. DNA methylation profiles (beta-values) for dendritic cells (n = 6; GEO accession GSE71837^66^) and hematopoietic stem cells (n = 5; GEO accession GSE63409^67^) were download using GEOquery (v2.68.0).

Data sets were combined (including 45 BPDCN samples), probes matching to chromosome X or Y were removed, and only probes present in all studies were to remove unwanted variation between the data sets (batch effects). Unwanted technical variation was removed using a two-stage approach (RUVm). First, a standard limma analysis was performed to identify empirical control probes (ECPs). Next, the results from stage 1 were used to perform a second differential DNA methylation with RUV-4 (*RUVfit* function provided by the MISSMETHYL package) and adjusted beta-values were extracted. Region-level analysis was performed to call differentially methylated regions (DMRs) using adjusted beta-values between BPDCN and dendritic cells using DMRcate (v2.13.0) and gene set testing was performed on DMRs with fdr < 0.1 and absolute difference above 0.3 against REACTOME and HALLMARK gene sets using the *gsaregion* function (MISSMETHYL). Linkage of genes and enriched REACTOME pathways was performed for DMRs with fdr < 0.01 and absolute difference above 0.3 using *enrichPathway* (REACTOMEPA, v1.44.0, *q*-value < 0.1) and *cnetplot* (ENRICHPLOT, v1.20.0).

### Comparative analysis with genome-wide DNA methylation data from acute myeloid leukemia

Regarding epigenetic discrepancies that may aid in the distinction of borderline cases between BPDCN and its predominant differential diagnosis, acute myeloid leukemia (AML), we comparatively analyzed our dataset with a comprehensive, previously published AML cohort, incorporating 243 samples for which genetic subtype and genome-wide DNA methylation data was available (GEO accession GSE159907^68^). Data (IDAT files) was processed (including 45 BPDCN samples) as described above using SWAN to normalize the data and probes from chromosome X or Y were removed. Unwanted variation was removed using a two-stage approach (RUVm). First, Illumina negative control (INCs) data for EPIC arrays was extracted (411 probes) and differential DNA methylation analysis using RUV-inverse with INCs as negative control features was performed. Next, the results from stage 1 were used to perform a second differential DNA methylation with RUV-4 and adjusted M-values were extracted for further analysis. Region-level analysis was performed to call differentially methylated regions (DMRs) using adjusted M-values between BPDCN and AML using DMRcate (v2.13.0) and gene set testing was performed on DMRs with fdr < 0.1 and absolute difference above 0.2 against REACTOME and HALLMARK gene sets using the *gsaregion* function (MISSMETHYL). Gene sets with *P* < 0.01 were considered as significantly enriched.

Linkage of genes and enriched REACTOME pathways was performed for DMRs with fdr < 0.01 and absolute difference above 0.2 using *enrichPathway* (*q*-value < 0.1) and *cnetplot*.

### Comparative analysis with genome-wide DNA methylation data from other entities

Raw data files (IDAT) from AML (n = 316; GEO accession GSE159907), T-cell acute lymphoblastic leukemia (t-ALL, n = 156; GEO accession GSE155339^69^), and melanoma (n = 450, TCGA project TCGA-SKCM) were downloaded and processed (including 45 BPDCN samples) as described above using SWAN to normalize the data; probes from chromosome X or Y were removed and beta-values were extracted. Additionally, beta-values for chronic myelomonocytic leukemia (CMML, n = 65; GEO accession GSE105420^24^) were downloaded and merged with the processed data. Unwanted variation was removed using a 2-stage approach as described in the section above (‘Comparative analysis with genome-wide DNA methylation data from different cell types’).

### Annotation of Methylated Regions

Probe annotations were extracted using the R package ILLUMINAHUMANMETHYLATIONEPICANNO.ILM10B4.HG19 (v0.6.0). Probes annotated as ‘Island’ were classified as CpG island (CGI) probes. Probes falling into 5’ UTRs or within 1,500 bp of the transcription start site were classified as promotor sites; sites within gene bodies or 3’UTRs were defined as gene body sites.

### Analysis of the tumor microenvironment by MethylCIBERSORT and immunohistochemistry

The cellular composition of the tumor microenvironment (TME) was assessed using MethylCIBERSORT as described. Beta values from raw IDATs and signature genes were deconvoluted according to immune cell populations ^18^. Partitioning around medoids (PAM) was applied to identify immunologically hot versus cold immune tumors based on MethylCIBERSORT calculations and to assign the optimal cluster number in the data (testing from 2 to 10). Results obtained by MethylCIBERSORT were then validated through a correlative immunohistochemical assessment (IHC) of the T-cell and monocyte fraction of the TME. Antibodies and positivity cut-offs employed in the current study remain as described ^6^.

### DNA-methylation-based mitotic clock estimation

To better comprehend the interaction between biological aging and molecular profiling in the light of the methylome, mitotic activity was estimated using epiCMIT. Mitotic clock was estimated on batch corrected beta-values for each sample and results were summarized per cohort ^27, 70^.

### FFPE-ATAC-seq

ATAC-sequencing on FFPE tissue sections from four typical pDC-like BPDCN patients was performed as described ^71^. Briefly, for nuclei isolation 20-µm-thick sections were deparaffined and underwent subsequent enzyme digestion. Then, 50.000 isolated FFPE nuclei were used in each FFPE-ATAC reaction composed of Tn5-mediated transposition and T7 in vitro transcription. FFPE-ATAC libraries were then sequenced on an Illumina NovaSeq 6000 platform at Novogene (Cambridge, UK) to a depth of at least 40 million 150-bp single-end or paired-end sequencing reads per library.

### Statistical analysis

If not reported otherwise, statistical analysis was performed using R (v4.3.0) and p-values were corrected using Benjamini-Hochberg correction. The following R packages were used: TIDYVERSE (v2.0.0)^51^ for data handling and plotting; MAFTOOLS (v2.17.0)^72^ to summarize, analyze, and visualize variant data; EnhancedVolcano (v1.18.0) (https://github.com/kevinblighe/EnhancedVolcano) plot volcano plots; COMPLEXHEATMAP (v2.16.0) and PHEATMAP (v1.0.12) to draw heatmaps; GGPUBR (v0.6.0) for box and violin-plots.

Intra-tumor heterogeneity was estimated on the entropy of somatic mutation (mDITHER-score) using DITHER (V1.0) ^73^. Correlations were calculated using Spearman’s rank correlation, if not stated differently.

## Data availability

Raw data for BPDCN samples have been downloaded from European Genome-phenome archive (EGA) under the previous accession number EGAS00001006166, additional cases were deposited under accession number EGAS00001007201, respectively. EPIC array data have been deposited in Gene Expression Omnibus (GEO) under accession number GSE230487.

## Pseudonymization

Processing of personal data for this study was performed pseudonymously by using a case-ID. Due to pseudonymization data backtracking specific to the individual is impossible. Only the initiators of the study (NG. AK, HW, JS) have access to a file that is separately stored (password-protected) containing the details on pseudonymization.

## Declarations

### Ethics approval and consent to participate

This retrospective study was approved by the ethics committee of the University of Lübeck (reference-no 18-311) and conducted in accordance with the declaration of Helsinki. Patients at the Reference center for Hematopathology have provided written informed consent regarding routine diagnostic and academic assessment, including genomic studies of their biopsy specimen alongside transfer of their clinical data.

## Competing interests

The authors declare that they have no conflict of interest.

## Funding

This work was supported by generous funding by the Stefan Morsch Foundation through a project grant (NG & HW). X.C. is supported by Swedish Research Council (2022-00658) and Swedish Cancer Foundation (21 1449Pj and 22 0491).

## Author contributions

Study concept: NG, ACF, HM

Data collection: NG, AK, JS, HW, SS, PL, KN, KK, VB, HM, SS, XC, PX

Data analysis and creation of figures and tables: AK, NG, HW, VB, HB, NvB, MS, XC, PX, FJ, HS

Initial Draft of manuscript: NG.

Critical revision and approval of final version: all authors.

## Acknowledgements

The authors would like to thank Tanja Oeltermann for her skilled technical assistance. AK and HB acknowledge computational support from the OMICS compute cluster at the University of Lübeck.

## Supplementary Figures

**Supplementary Figure 1**: Deconvolution of bulk RNA-seq from the extended cohort of BPDCN patients.

**Supplementary Figure 2**: Complete representation of promotor and gene body DNA methylation of TSGs (according to Vogelstein *et al.*).

**Supplementary Figure 3**: Mutational landscape according to BPDCN C1 vs C2. a significantly differentially mutated genes between C1 and C2. b TMB according to C1 vs C2

**Supplementary Figure 4**: Correlating the results from our MethylCIBERSORT approach with the genomic landscape

**Supplementary Figure 5**: Mutational, epigenetic and immunohistochemical properties according to ICs

**Supplementary Figure 6**: Quantitative estimation of tumor-infiltrating T-cells by **a** IHC and **b** MethylCIBERSORT

**Supplementary Figure 7**: Unsupervised clustering of most variable methylated probes (n = 5,600) a. Heatmap b. TMB according to MethCs

## Supplementary Tables

**Supplementary Table 1: Variants**

**Supplementary Table 2: MutSigCV**

**Supplementary Table 3: Clinical data**

**Supplementary Table 4: beta values for differentially methylated probes in the BPDCN cohort.**

**Supplementary Table 5: a) Correlation between DNA methylation and gene expression for selected Genes (TSGs and oncogenes). b) Correlation between DNA methylation and gene expression for the entire transcriptome.**

## Notes

**Competing Interests:** The authors declare no conflicts of interest.

### Competing Interest Statement

The authors have declared no competing interest.

### Author Declarations

This retrospective study was approved by the ethics committee of the University of Luebeck (reference-no 18-311) and conducted in accordance with the declaration of Helsinki. Patients at the Reference center for Hematopathology have provided written informed consent regarding routine diagnostic and academic assessment, including genomic studies of their biopsy specimen alongside transfer of their clinical data.

