## Supplementary figures and images for "Genome-wide DNA methylation-analysis delineates blastic plasmacytoid dendritic cell neoplasm from related entities and identifies distinct molecular features"

### Supplementary Figure 1

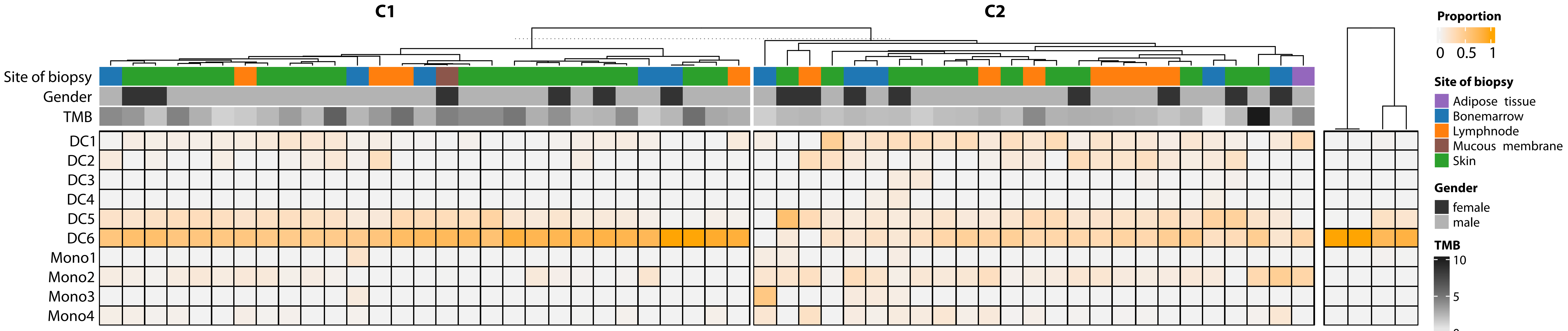

### Supplementary Figure 2

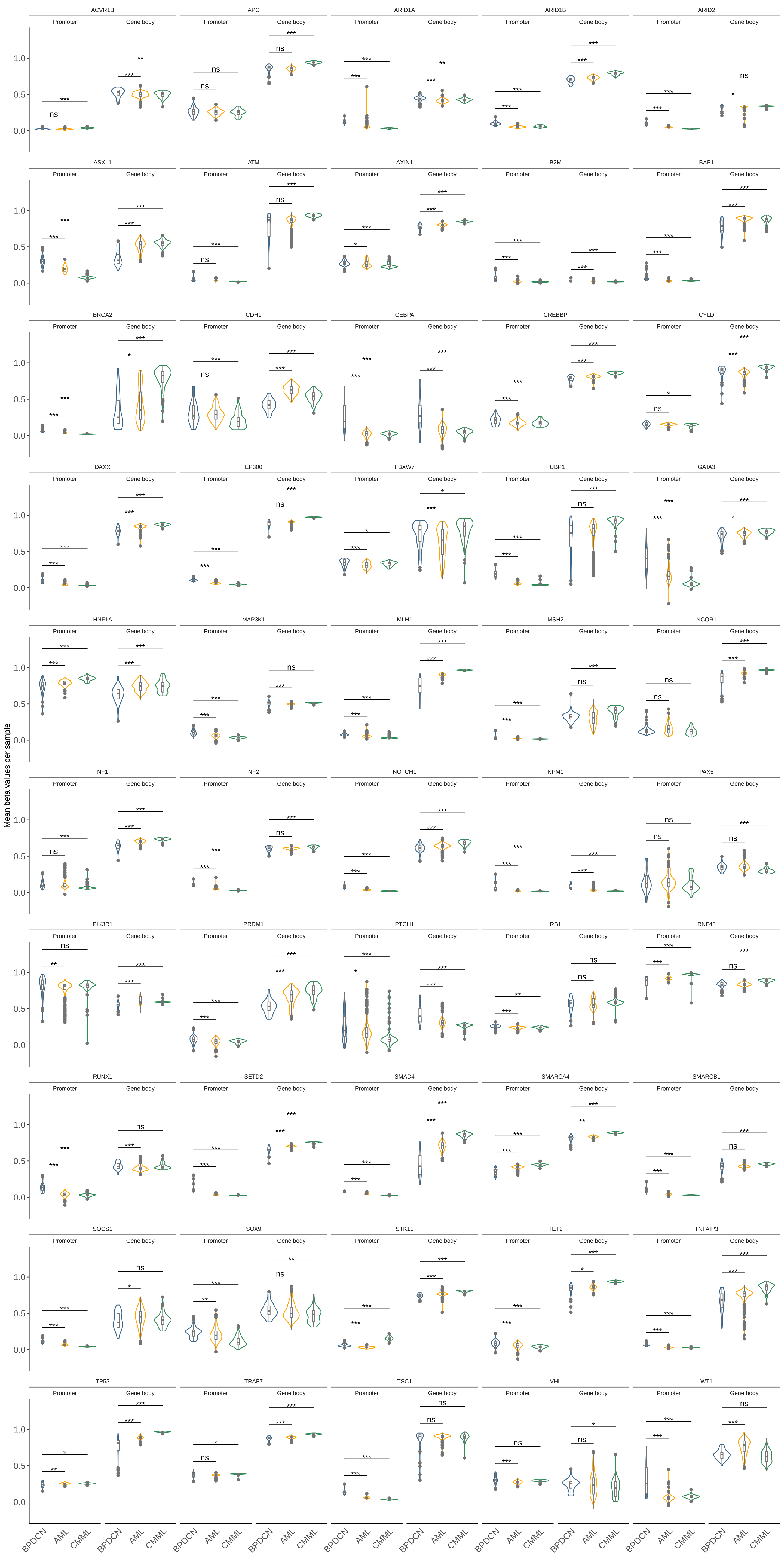

### Supplementary Figure 3

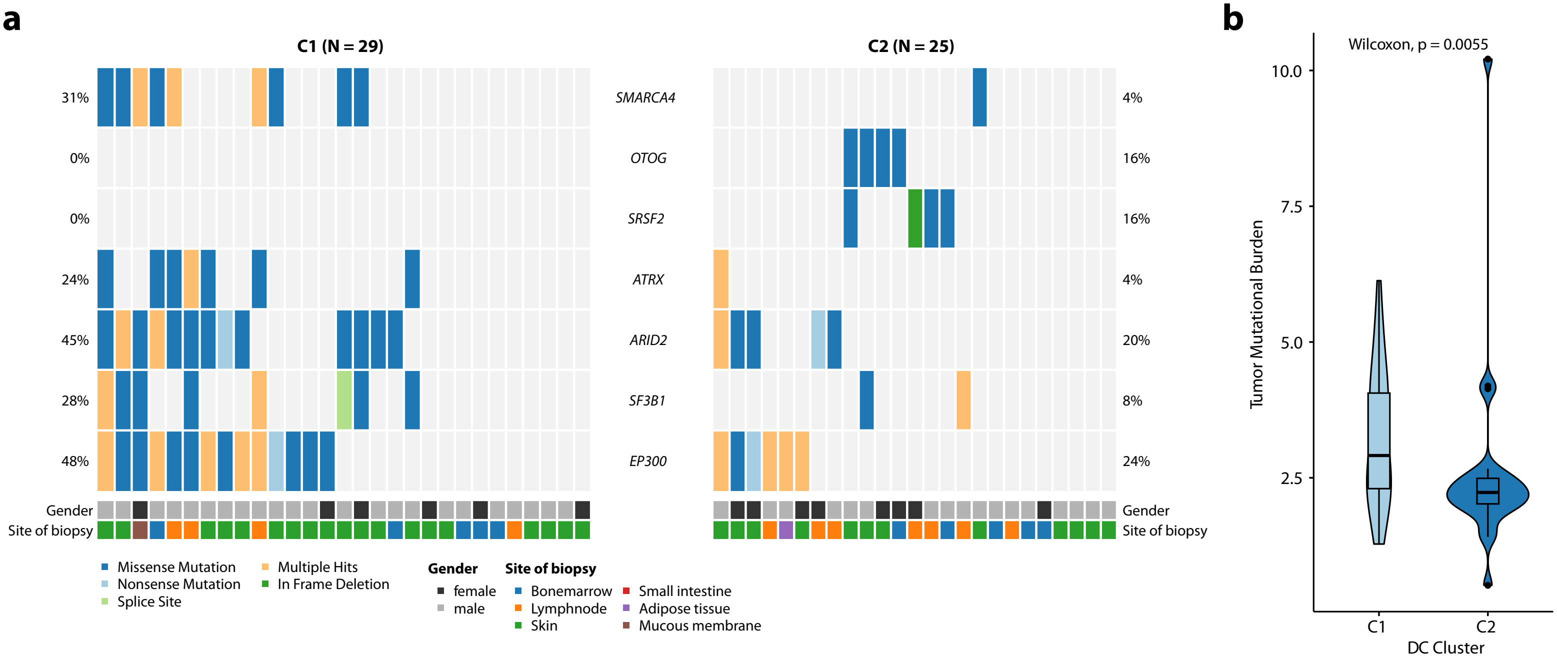

### Supplementary Figure 4

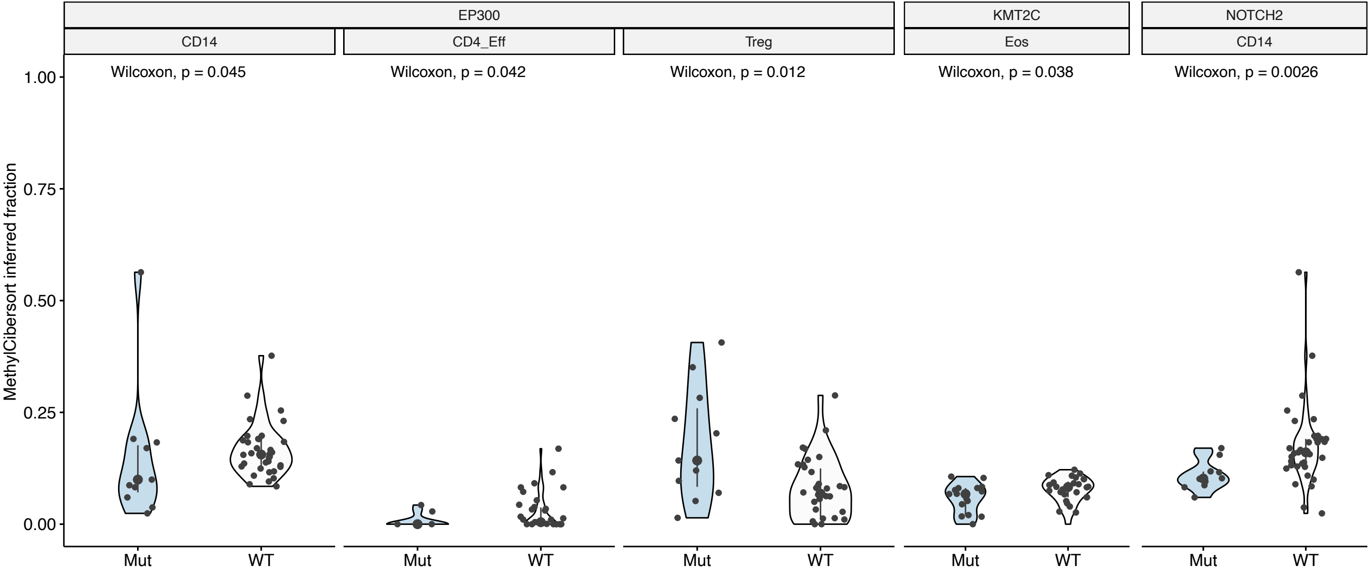

### Supplementary Figure 5

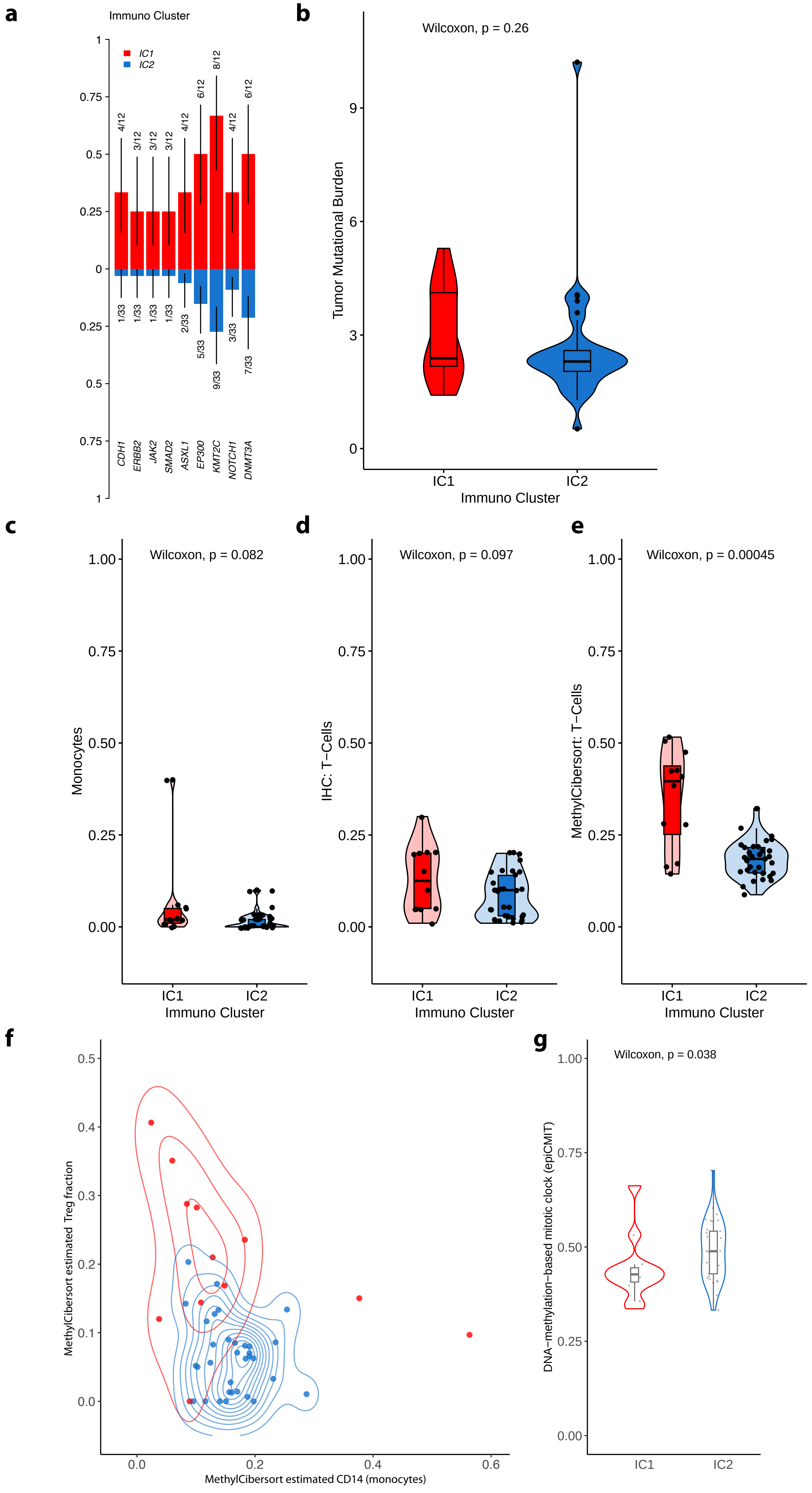

### Supplementary Figure 6

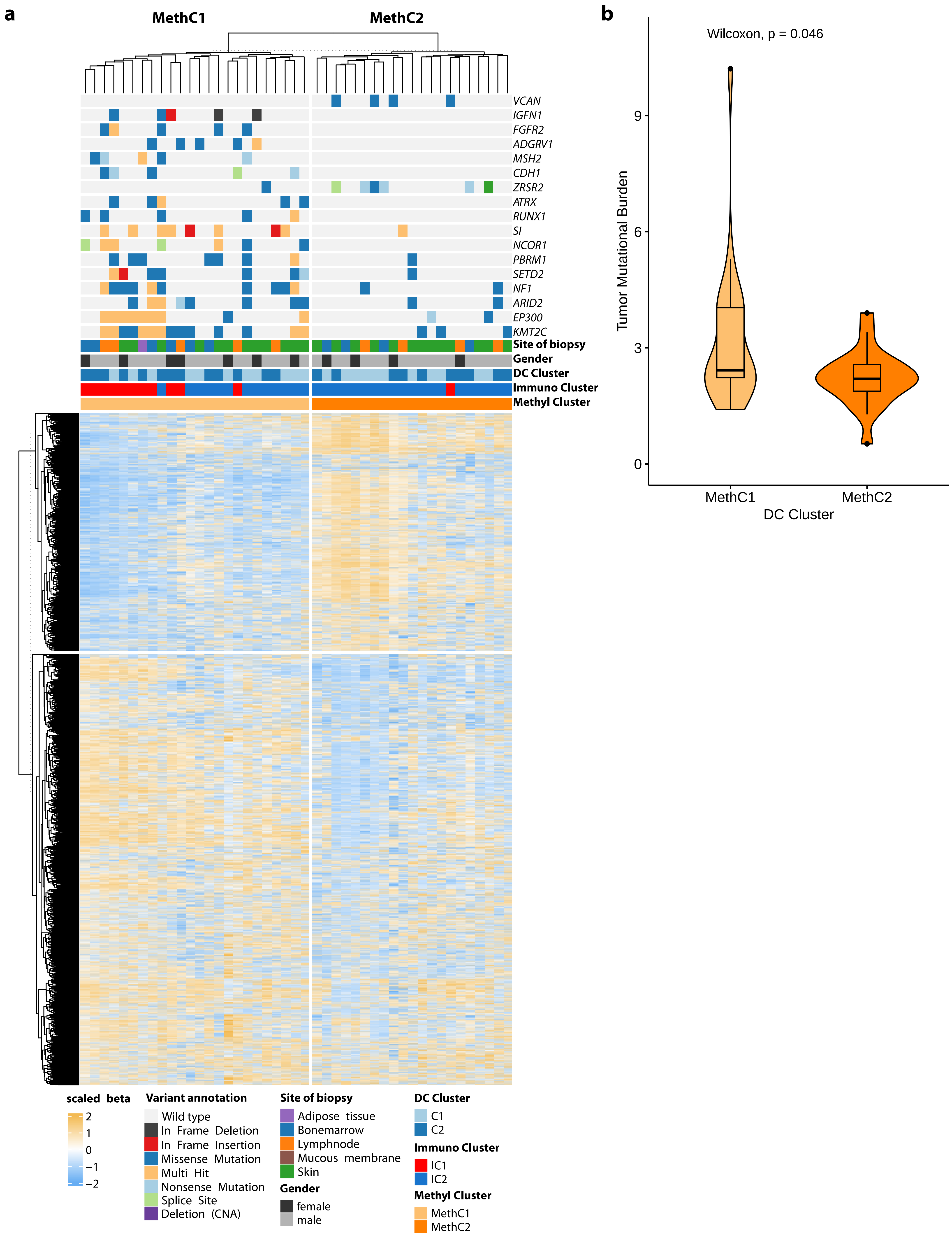

### Supplementary Figure 7

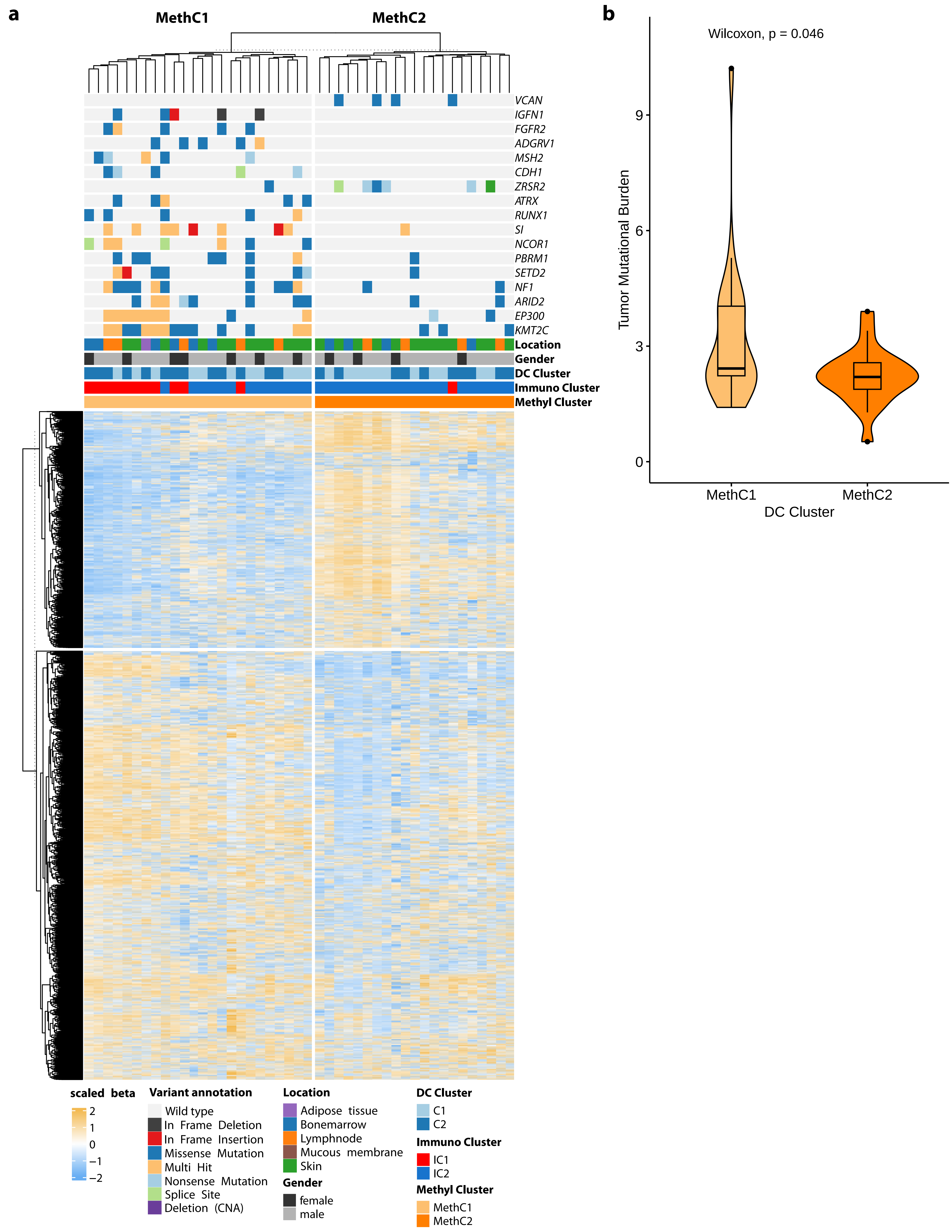
